# Repurposing chlorpromazine as add-on in the adjuvant phase of first-line glioblastoma therapeutic protocol in patients carrying hypo-/un-methylated *MGMT* gene promoter: RACTAC, a Phase II multicenter single-arm clinical trial

**DOI:** 10.1101/2023.02.21.23286088

**Authors:** Andrea Pace, Giuseppe Lombardi, Veronica Villani, Dario Benincasa, Claudia Abbruzzese, Ilaria Cestonaro, Martina Corrà, Giulia Cerretti, Mario Caccese, Antonio Silvani, Paola Gaviani, Diana Giannarelli, Marco G. Paggi

**Author notes:** **Corresponding author:** Marco G. Paggi, MD, PhD IRCCS - Regina Elena National Cancer Institute Via Elio Chianesi 53, 00144 Rome, Italy.

## Abstract

**Background:** Glioblastoma (GBM) is a devastating brain tumor with poor prognosis, characterized by rapid growth and invasion into surrounding brain tissue. It is a hard-to-treat cancer and represents an unmet medical need. In recent years, there has been a growing interest in developing novel approaches to improve the outcomes of GBM patients; among these, drug repurposing. Our preclinical studies identified the antipsychotic chlorpromazine (CPZ) as an important modulator of signal transduction and energy metabolism in GBM cells, so we embarked on a Phase II clinical trial in which CPZ has been added to the standard disease treatment.

**Methods:** With these assumptions, we started a multicenter phase II clinical trial on newly diagnosed GBM patients carrying hypo-/un-methylated *MGMT* gene promoter by adding CPZ to temozolomide (TMZ) in the adjuvant phase of the standard first-line therapeutic protocol RACTAC schedule). Primary endpoints: Progression-Free Survival (PFS) and Combination treatment toxicity. Secondary endpoints: Overall Survival (OS) and Quality of Life (QoL)

**Results:** The RACTAC schedule showed an overall clinical benefit in GBM patients carrying hypo-/un-methylated *MGMT* gene promoter. When compared with historical cohorts, these patients displayed longer PFS, with toxicity described as a dose-dependent sedation and liver toxicity, both expected. One case of severe liver toxicity has been reported. OS and QoL are still under evaluation.

**Conclusions:** This clinical trial confirms the anticancer properties of CPZ, as described in several preclinical studies. In addition, the RACTAC study can be considered at least as a proof-of-concept in demonstrating the effectiveness of interfering with the well-described oncogenic monoamine signaling between neurons and GBM.

## Introduction

Glioblastoma (GBM) is the most frequent and severe brain tumor, characterized by poor response to treatment and an almost certainty of relapse. First-line GBM treatment, regardless of the molecular classification of the disease ^1^, consists in maximal surgical resection followed by radiotherapy with concomitant temozolomide (TMZ) treatment, followed by adjuvant TMZ administration. This scheme is however associated with a median overall survival of 14.6 months and a 5-year survival <5% ^2^, a clinical course that clearly denotes an unmet medical need.

Given its peculiar biological characteristics, GBM presents high invasive and infiltrative properties ^3^, also coupled with a distinctive cellular heterogeneity, a prerequisite for a swift adaptation under therapeutic pressure ^4–7^. A key role in GBM resistance to therapy is played by its ability to recover from genetic damages induced by radiotherapy and TMZ, by means of an effective DNA repair, especially in tumors characterized by unmethylated O6-methylguanine methyltransferase (*MGMT*) gene promoter ^8^. Of note, GBM is among the few tumors in which a single-drug treatment is currently in use, in contrast with most therapeutic approaches in oncology and with its intratumor heterogeneity.

An interesting characteristic of GBM is its responsiveness to neurotransmitters, as monoamines^9–11^. The well-known interplay between neurons and tumors, especially GBM ^10, 12^, and the more recent identification of a synaptic neuron-GBM connectivity confirms that neuron-secreted mediators are taken up by GBM cells, where they act as oncogenic stimuli ^13^. These findings paved the way for considering the addition of selected neuroleptic drugs as potentially effective addition to GBM treatment ^14^. Indeed, Many psychotherapeutic drugs, act on several post-synaptic receptors and display polypharmacological activity ^14^.

With these assumptions, we chose to evaluate the effects of one of the progenitors of neuroleptic medications, i.e., the antipsychotic drug chlorpromazine (CPZ), in use for about 70 years for the therapy of several psychiatric disorders. CPZ is a compound listed in the 2021 WHO Model List of Essential Medicines^15^.

Delving into CPZ pharmacodynamics *in vitro*, our group assayed its effects on established and primary human GBM cells. The results defined the role of this drug in hindering GBM cell growth by acting at different levels through multimodal antitumor effects without any action on non-cancer neuroepithelial cells^16, 17^. In addition, CPZ acts synergistically with TMZ in hindering GBM vitality and stemness capabilities ^16^.

CPZ, besides not devoid of side effects, is a safe, low-cost and promptly available medication. All these conditions paved the way for repurposing CPZ as an add-on drug in GBM therapy. To this end, we planned the RACTAC (<u>R</u>epurposing the <u>A</u>ntipsychotic drug <u>Ch</u>lorpromazine as a <u>T</u>herapeutic <u>A</u>gent in the <u>C</u>ombined treatment of newly diagnosed glioblastoma) Phase II multicenter single arm study, in which CPZ is added to adjuvant TMZ in the first-line therapy of GBM patients whose tumor is characterized by an unmethylated *MGMT* gene promoter, i.e., those characterized by resistance to TMZ and overall poorer prognosis ^8^.

## Patients and Methods

### Study design

This was a multicenter, prospective, single-arm clinical trial. The experimental procedure involved the combination of CPZ with standard treatment with TMZ in the adjuvant phase of the Stupp protocol ^2^ in newly diagnosed GBM patients carrying an unmethylated *MGMT* gene promoter. All patients received CPZ (“Largactil”, from Teofarma S.R.L., Valle Salimbene (PV), Italy, as 25 mg tablets during the whole period of TMZ adjuvant treatment at a starting dose of 25 mg/day orally from day 1 of the TMZ adjuvant chemotherapy. The dosage was increased to 50 mg/day at the second cycle of treatment, if well tolerated, and continued for 6 cycles or until disease progression, death, unacceptable toxicity, or consent withdrawal (**Figure 1**).

**Figure 1.**
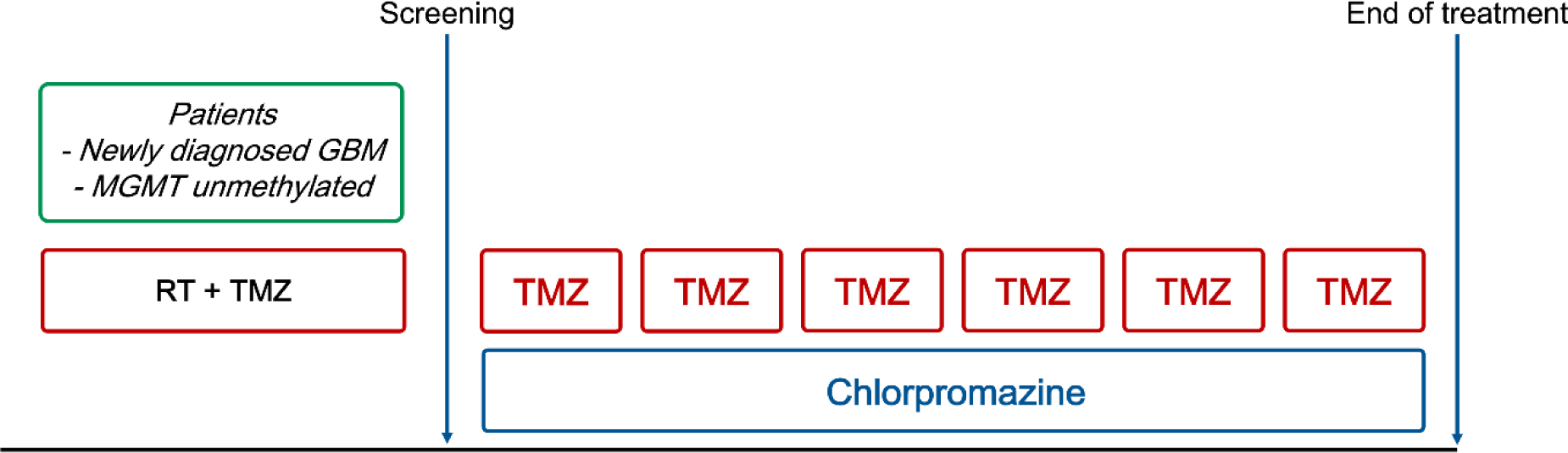
Scheme of the daily addition for of CPZ during the adjuvant phase of the first line protocol for newly diagnosed GBM patients.

This Phase II clinical trial has been approved by our Institutional Ethics Committee (Comitato Etico Centrale IRCCS - Sezione IFO-Fondazione Bietti, Rome, Italy) on September 6, 2019.

The study is registered as EudraCT #2019-001988-75 and ClinicalTrials.gov Identifier #NCT0422444. All patients provided informed written consent prior to participating in any study procedures or treatments.

### Eligibility

Patients aged 18-75 years with newly diagnosed and histologically confirmed supratentorial GBM (World Health Organization grade 4) and unmethylated *MGMT* gene promoter status were eligible. Diagnosis of GBM was based on neurosurgical resection of the tumor or biopsy. Additional inclusion criteria were: age ≥18 years; gadolinium-enhanced MRI, 48 h after surgery or before randomization; stable or decreasing dose of steroids for 1 week before randomization; Karnowski performance status(KPS) >70; satisfactory laboratory blood and biochemical results within 2 weeks prior to randomization. Local assessment of O-6-methylguanine-DNA methyltransferase (MGMT) methylation status on tumoral tissue was required.

### Endpoints

Progression-Free Survival (PFS) was the primary endpoint. PFS was defined as the time from the start of the Stupp regimen (radiotherapy plus concomitant TMZ) to the earliest documented date of disease progression, based on Response Assessment in Neuro-Oncology (RANO) criteria ^18, 19^, as determined by the investigator, death due to any cause, or censored at the date of the last assessment. Evaluating a meta-analysis performed on 91 GBM clinical trials, the choice of PFS as an endpoint can be considered appropriate as a surrogate endpoint for earlier evaluation. The 6-mos PFS appears correlated with 1-year overall survival (OS) and median OS ^20^.

Secondary endpoints were as follows: (i)Overall survival (OS; defined as the time from diagnosis to the date of death from any cause or loss-to-follow-up; (ii) combination treatment toxicity; adverse events (AEs) were evaluated according to the National Cancer Institute’s Common Terminology Criteria for Adverse Events version 5.0. Safety assessments were performed from treatment initiation through 30 days after the last dose of chemotherapy with TMZ plus CPZ; (iii) Quality of Life (QoL), assessed by means of the EORTC QLQ C30+BN20 questionnaire at baseline and every 3 cycles ^21, 22^.

### Follow-up

Patients were followed-up monthly during adjuvant chemotherapy, and every 3 months. after completion of the standard treatment if the disease was stable. Radiological assessment was done by gadolinium (Gd) brain MRI every 12 weeks from the first drug administration until disease progression.

Response assessment during study was based on investigators’ evaluation using the RANO criteria ^18, 19^, which included blinded MRI readings, assessment of neurological status, KPS scores, and steroid use.

### Statistical analysis

All analyses were performed on the patients who received at least one dose of the study treatment. The primary objective of the study was to evaluate the proportion of patients free from progression after 6 months (PFS-6). Considering as unacceptable a percentage of PFS-6 (P0) equal to 35% and a desirable PFS-6 of 55% (P1), a minimum of 41 patients would be needed to guarantee a power of 80% at a significance level of 5% (one-sided), according to A’Hern ^23^. If at least 20 patients were progression-free after 6 months, the treatment would be considered sufficiently active. All survival curves were estimated by the Kaplan-Meier method.

Clinical and demographic characteristics are reported as absolute counts and percentage when related to categorical items and as median and range if referred to quantitative variables.

## Results

### Patient characteristics

Between April 2020 and August 2022, 45 patients who met the eligibility requirements and consented to participate were screened. Forty-one patients received at least one cycle of treatment and were included in the final analysis. Study follow-up was closed on December 31, 2022. Patients’ baseline characteristics and demographics are summarized in **Table 1**.

**Table 1.** Baseline characteristics of the enrolled GBM patients.

|  | **Baseline characteristics** |
| --- | --- |
| Variables | Study population (n= 41) |
| Age at diagnosis<br>(Median, y) | 56 (range 20-75) |
| Gender | M 29 (70.7%) |
| KPS median | 90 (range 70-100) |
| Extent of resection % | GTR 41.5; Partial 56.1; Biopsy 2.4 |
| Steroid treatment at<br>baseline # (%) | 21 (51%) |
| Dexamethasone dose<br>(mean) | 4 mg |

KPS=Karnofsky Performance status; GTR=Gross Total resection

Twenty patients (48,7%) completed 6 cycles of treatment with TMZ at standard dose plus CPZ; twenty-one patients (51.2%) discontinued the treatment due to early progression (mean cycles completed = 4.6). The median follow-up of was 15 months. (range 3–37).

### Progression-free survival

At 6 months, 27 patients were alive and without progression thus making possible the achievement of the primary endpoint. Overall, 35 patients (85.4%) underwent disease progression, with a median PFS of 8.0 months. (95% CI: 7.0-9.0) (**Figure 2A**). PFS at different times is reported in **Table 2**.

**Figure 2.**
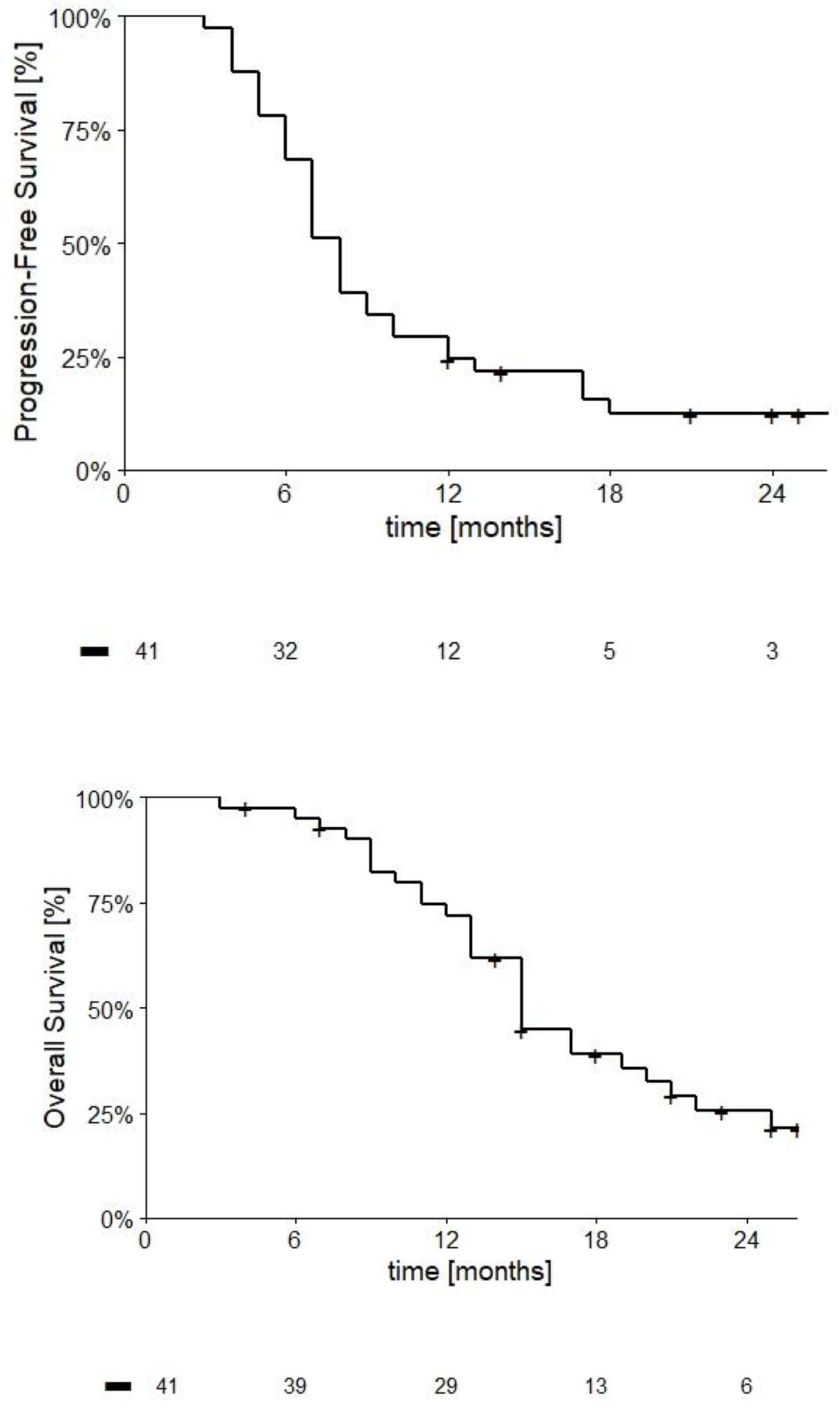
Kaplan-Meier curves describing PFS and OS trends in GBM patients with unmethylated *MGMT* gene promoter treated according to the RACTAC protocol.

**Table 2.** Progression-free survival (PFS) and overall survival (OS) in GBM patients with unmethylated *MGMT* gene promoter treated according to the RACTAC protocol.

|  | 6 months | 12 months | 18 months | 24 months |
| --- | --- | --- | --- | --- |
| PFS | 68.3% | 24.4% | 12.4% | 12.4% |
| OS | 95.1% | 72.0% | 38.9% | 25.5% |

### Overall survival

At the date of study closure, 29 deaths (69.7%) were observed, and median overall survival has been 15.0 months (95% CI: 13.1-16.9). OS at different time-points is reported in **Figure 2B** and **Table 2**.

### Toxicity

. The RACTAC clinical experimentation has been correlated with the onset of an expected dose-dependent sedation, especially at the beginning of the CPZ administration, and liver toxicity (1 serious case), however expected for both CPZ and TMZ. Treatment was well tolerated in all patients with mild somnolence (grade 1-2) in 8 (19,5%) and asthenia in 12 (29,2%) patients. In three patients, grade 3-4 hyper-transaminasaemia was observed (7%). Adverse events lead to reduction or interruption of CPZ dosage in 4 patients (9,7%).

Decreased platelets count grade 3 was observed in one patient and neutropenia grade 3 in one. All these data are summarized in **Table 3**.

**Table 3.** Drug-related adverse events.

| Adverse events | Grade 1-2 | Grade >2 |
| --- | --- | --- |
| Somnolence | 8 | - |
| Fatigue | 11 | 1 |
| Hypertransaminasemia | 1 | 2 |
| Thrombocytopenia | 6 | 1 |
| Neutropenia | 1 | 1 |

## Discussion

GBM patients with unmethylated *MGMT* promoter gene comprise 55-60% of total GBM cases and present poorer prognosis due to an intrinsic resistance towards therapy with alkylating agents. At present, while it is well recognized that TMZ brings a benefit in patients with methylated *MGMT* gene promoter, it remains controversial whether this medication should be administered or omitted when treating GBM patients with unmethylated *MGMT* gene promoter. Indeed, EANO evidence-based guidelines on diagnosis and treatment of diffuse gliomas of adulthood report that TMZ might only be active in GBM patients with methylated *MGMT* gene promoter, whereas its effectiveness on patients with unmethylated *MGMT* gene promoter appears modest ^8, 24^.

Various therapeutic strategies have been investigated in several Phase II clinical trials, with the aim to overcome drug resistance or to overpass DNA repair pathways in GBM patients with unmethylated *MGMT* gene promoter ^8, 25^. In the same setting, addition of drugs other than TMZ alone did not lead to encouraging results ^26–28^. Other studies evaluated the efficacy of new agents in replacement of TMZ in the first line treatment of GBM patients with unmethylated *MGMT* gene promoter. In the EORTC 26082 Phase II trial, temsirolimus was administered either during radiotherapy or as adjuvant treatment, showing no clinical benefit ^29^. In a single arm phase II trial, enzastaurin, a protein kinase C inhibitor, was administered before, concomitantly with, and after radiotherapy to determine PFS-6 months in GBM patients with unmethylated *MGMT* gene promoter, not achieving the primary endpoint ^30^. On the other hand, in the same setting, therapy with bevacizumab plus irinotecan provides a PFS at 6 months better than TMZ, but with no improvement in OS ^31^.

Our results show that the addition of CPZ to standard TMZ in the first line treatment of GBM patients with unmethylated *MGMT* gene promoter was safe and led to a longer PFS than the one expected in this population of patients. A recent meta-analysis of pooled data from five Phase III clinical trials reported PFS and OS a in patients with unmethylated *MGMT* gene promoter of 4.99 and 14.11 months, respectively, when the standard-of-care GBM treatment was used ^32^.

In our study, however, median OS was 15 months, thus not representing a clinically-relevant improvement with respect to data reported in this meta-analysis. Such an outcome could also relate to the administration of CPZ only in the adjuvant phase of treatment. Further studies are needed to test the concomitant association of CPZ also during radiotherapy. The safety profile of CPZ was consistent with its well-known pharmacological profile, even when administered concomitantly with TMZ. In our study, the most frequent adverse event were mild somnolence (grade 1-2) in 18% of patients, usually in the first month of treatment, and fatigue, observed in 30% of patients, a symptom commonly reported within a range of 40-70% in primary brain tumor patients^33^.

Quality of life (QoL) is an important outcome to be evaluated in these GBM patients. It has been measured, as specified above, by means of the EORTC QLQ-C30 and QLQ-BN20 questionnaires. Presently, these evaluations are in progress and the results will be published shortly.

Most relevant limitations of RACTAC trial are the small number of patients included in the study and the lack of a control group. However, the clinical characteristics of patients at baseline seem to be representative of the real-world population of MGMT unmethylated GBM. PFS6 observed in this trial was 8 months and resulted promisingly longer than that reported in previous published trials in this population.

Since the plasticity of GBM and its ability to remodulate its cell population based on the selective pressure generated by therapies, we can state that this tumor cannot be defined as a “single-path disease”, being therefore very unsatisfying to treat it by means of targeted therapies. On these premises, it appears reasonable to consider the opportunity to use “dirty drugs”, i.e., drugs that are not too targeted, but are able to hit some generalized vulnerabilities characterizing cancer cells.

CPZ is a well-known DRD2 antagonist ^34^ and therefore has been successfully used in the treatment of psychiatric disorders. We intended to take advantage of its ability to interfere with the function of DRD2, as well as a number of other neuromediator receptors (https://go.drugbank.com/drugs/DB00477), to hamper the pseudo-synaptic, oncogenic interplay between neurons and GBM.

Indeed, the results of the RACTAC clinical trial reported here can be considered at least as proof-of-concept in supporting the effectiveness of interfering with the oncogenic monoamine signaling between neurons and GBM. Of note, it should be considered that there is also a profound interplay between peripheral/central nervous systems and cancer, which acts through monoamine neuromediators and could represent a vulnerability targetable through the use of repurposed neuropsychiatric drugs in oncology and appliable to diverse cancers ^35, 36^. Evaluating the features of CPZ possibly useful in GBM treatment, we could also consider a number of studies showing the ability of this drug to hinder GBM malignant features in preclinical settings ^37^. In the same context, during the course of the RACTAC clinical trial, our group further refined the pharmacological effects of CPZ on GBM cells *in vitro*, demonstrating the ability of this medication to hinder GBM malignant features at multiple levels ^16, 17^ other than its known interference with the activity of neurotransmitters.

Considering the escalation of the costs of novel anticancer medications, the long time it takes for them to reach the market and the consequent nonavailability for a great number of patients, the use of repurposed drugs can dramatically cut down time and drug expenses for effective medications to reach the bedside, with significant benefits for the Health Systems. In addition, the characteristics inherent in repositionable drugs represent a further therapeutic chance for GBM patients for which no second-line therapy is currently established as effective or for those that have already experienced all known therapeutic opportunities.

## Data Availability

All data produced in the present study are available upon reasonable request to the authors.

## Acknowledgments

Work partially financed by Funds Ricerca Corrente 2018-2019, 2020-2021, 2022-2023 from Italian Ministry of Health (MGP) and by the European Union - NextGenerationEU through the Italian Ministry of University and Research under PNRR - M4C2-I1.3 Project PE_00000019 “HEAL ITALIA” to Marco G. Paggi, CUP H83C22000550006. The views and opinions expressed are those of the authors only and do not necessarily reflect those of the European Union or the European Commission. Neither the European Union nor the European Commission can be held responsible for them.

This paper is dedicated to the memory of our friend and colleague Armando Felsani, who recently passed away and who contributed so much to the advancement of knowledge in the field of cell differentiation and cell cycle regulation.

Editorial and graphical assistance were provided by Aashni Shah, Valentina Attanasio and Massimiliano Pianta (Polistudium SRL, Milan, Italy), and was supported by internal funds.

## Notes

### Competing Interest Statement

The authors have declared no competing interest.

### Clinical Trial

EudraCT # 2019-001988-75; ClinicalTrials.gov Identifier: NCT04224441

### Clinical Protocols

https://clinicaltrials.gov/ct2/show/record/NCT04224441

https://www.clinicaltrialsregister.eu/ctr-search/search?query=2019-001988-75

### Author Declarations

This study has been approved by the Institutional Ethics Committee (Comitato Etico Centrale IRCCS - Sezione IFO-Fondazione Bietti, Rome, Italy) on September 6, 2019

### Summary of Updates

Materials and Methods Results Discussion

